# Quantifying the rebound of influenza epidemics after relaxing nonpharmaceutical interventions during the coronavirus disease 2019 pandemic in China

**DOI:** 10.1101/2022.12.18.22283627

**Authors:** Hao Lei, Lei Yang, Mengya Yang, Jing Tang, Jiaying Yang, Minju Tan, Shigui Yang, Dayan Wang, Yuelong Shu

## Abstract

The co-existence of coronavirus disease 2019 (COVID-19) and seasonal influenza epidemics has become a potential threat to human health, particularly in China in the oncoming season. However, with the relaxation of nonpharmaceutical interventions (NPIs) during the COVID-19 pandemic, the rebound extent of the influenza activities is still poorly understood. In this study, we constructed a susceptible-vaccinated-infectious-recovered-susceptible (SVIRS) model to simulate influenza transmission and calibrated it using influenza surveillance data from 2018 to 2022. We projected the influenza transmission over the next 3 years using the SVIRS model. We observed that, in epidemiological year 2021–2022, the reproduction numbers of influenza in southern and northern China were reduced by 64.0% and 34.5%, respectively, compared with those before the pandemic. The percentage of people susceptible to influenza virus increased by 138.6% and 57.3% in southern and northern China by October 1, 2022, respectively. After relaxing NPIs, the potential accumulation of susceptibility to influenza infection may lead to a large-scale influenza outbreak in the year 2022-2023, the scale of which may be affected by the intensity of the NPIs. And later relaxation of NPIs in the year 2023 would not lead to much larger rebound of influenza activities in the year 2023-2024. To control the influenza epidemic to the pre-pandemic level after relaxing NPIs, the influenza vaccination rates in southern and northern China should increase to 53.8% and 33.8%, respectively. Vaccination for influenza should be advocated to reduce the potential reemergence of the influenza epidemic in the next few years.

## Introduction

In December 2019, the transmission of a novel coronavirus, severe acute respiratory syndrome coronavirus 2 (SARS-CoV-2), led to a pandemic of coronavirus disease 2019 (COVID-19) [1]. To mitigate the COVID-19 pandemic, a set of nonpharmaceutical interventions (NPIs), including massive mobility restrictions, universal fever screening, the use of big data and artificial intelligence to strengthen contact tracing, and the management of prioritized populations, were implemented worldwide [2]. Because both COVID-19 and seasonal influenza are respiratory infections and have similar routes of transmission [2][5], it is not surprising that influenza activity decreased under COVID-19 control measures during the 2019–2020 seasons worldwide, such as in Singapore [6], China [7][9], the United States [10], and New Zealand [11]. The impact of NPIs on COVID-19 transmission has been well-documented [12, 13]. However, long-term NPIs have raised concerns about the potential for economic recession [14] and unintended adverse mental health outcomes [15]. With more people being vaccinated against COVID-19, NPIs have been relaxed worldwide [16].

NPIs used to control COVID-19 significantly reduced seasonal influenza transmission at the beginning of the COVID-19 pandemic in 2020 [6,11]. However, their potential long-term impact is less well-understood, especially when NPIs are relaxed. A study had quantified the long-term impact of NPIs used to control COVID-19 on the influenza epidemic in the United States [17]. And a sharp increase in influenza activity during the 2021–2022 season has been reported in some countries in which NPIs were relaxed [18]. However, different to the situation in the United States and Europe, longer and stricter NPIs were implemented in China. Thus, influenza activity was at a low level during the past two seasons in China, and it is possible that human immunity to the influenza virus was also at a low level. Thus, with the relaxation of NPIs, it is possible that there was a sharper increase of influenza activity in the following seasons in China. In addition, with the relaxation of NPIs, the co-existence of coronavirus disease 2019 (COVID-19) and seasonal influenza epidemics in the oncoming season would become a potential threat to human health in China [19, 20].

In this study, we addressed these challenges by building a susceptible-vaccinated-infected-recovered-susceptible (SVIRS) model that was applied to influenza surveillance data in China from 2018 to 2022, and we used this model to generate predictions after 2022. This study aimed to quantify the long-term impact of NPIs on the influenza epidemic by predicting influenza transmission dynamics over the next 3 years under various control scenarios.

## Methods

### Data source

Weekly influenza surveillance data from 2018 to 2022 were obtained from the Chinese National Influenza Center. The influenza surveillance data reported each week included the number of hospital visits, influenza-like illness (ILI) cases, specimens tested, and number of positive influenza A (H1N1, H3N2, and pdmH1) and influenza B (Yamagata and Victoria) cases examined in the laboratory. The weekly incidence rate of influenza was calculated by multiplying the ILI rate of patients in the sentinel hospital by the positive rate of viral detection, and the weekly incidence rate was converted into the daily incidence rate using splines [21]. The daily incidence rates of influenza A (H1N1, H3N2, and pdmH1) and influenza B (Yamagata and Victoria) were combined to obtain the total daily incidence rate of influenza. Northern China and southern China experience different influenza seasonal patterns [22-24]. In addition, it’s possible that the NPIs implemented in the northern and southern China may be different due to the socioeconomic factors, so that the long-term impact of NPIs on influenza activities may be different. Therefore, in this study, influenza transmission in southern and northern China was studied separately. The epidemiological year of influenza is defined as being from October 1 of one year to September 30 of the following year, and this definition is based on the seasonality of the influenza epidemic in China [23, 24].

### SVIRS Model

We constructed a SVIRS model to simulate and project the influenza transmission in China. The governing equations are as follows:

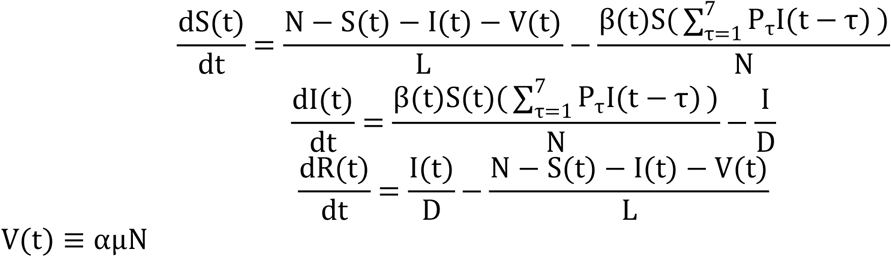

where N represents the total population in northern China or that in southern China; S(t), I(t), R(t) and V(t) represent the numbers of susceptible, infected, recovered, and vaccinated individuals, respectively, at time t; N=S(t)+I(t)+R(t)+V(t), L indicates the average duration of immunity, and D is the mean infectious period. μ represents the influenza vaccination rate, α represents the vaccine effectiveness, and α = 59% (95% CI: 51.0%–66.0%) [25]. β(t) is the transmission efficiency of the influenza virus at time t. As climate can influence influenza transmission, it was assumed, for simplicity, that β t = a(1+bsin(wt+c)) [26], where b represents the relative contribution of climate to the spread of influenza, which generally ranges from 0.05 to 0.3 [26]. In addition, P_τ_ is the 7-day infectivity profile, which is subject to a gamma distribution with a mean of 2.7 days and a variance of 1.8 days [27]. i(t) is the number of new infected cases at time t, and 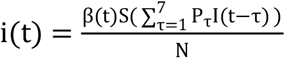. rate(t) represents the influenza incidence rate at time t, and 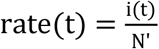, where N’ indicates the reporting population of influenza cases, and we assumed that N’ = 10%N [26]. According to previous studies, w was assumed to be 0.017(=2π/365), which means that β t is a periodic function of 1 year [28]. D was fixed at 5 days, and L was fixed at 2 or 3 years [29] to explore how the incidence rate of influenza will change after relaxing various NPIs.

### Initial condition

The initial percentage of infected people, I(0), in each epidemiological year was chosen as the 40% quantile of the nonzero weekly incidence rate. The timing of the onset of the influenza epidemic is always defined as the first 3 consecutive weeks with a weekly incidence rate exceeding the 40% quantile of the nonzero weekly incidence rate [29, 30]. The initial percentage of susceptible individuals is always unknown because of several factors that are unknown, such as the influenza virus evaluation and dominant strain in that year. In this study, the initial percentage of susceptible individuals, S(0), in epidemiological year 2018–2019 was estimated based on the assumption that the simulated percentage of susceptible individuals at the end of epidemiological year 2018–2019 was equal to S(0) in that epidemiological year. This assumption is based on the idea that, in the years before the COVID-19 pandemic, the influenza epidemic was relatively stable; therefore, in each epidemiological year, the initial percentage of susceptible individuals was the same, and the influenza transmission dynamics in epidemiological year 2018–2019 were similar to those in epidemiological year 2019– 2020. The unknown parameters a, b, and c in the model were estimated using the Markov Chain Monte Carlo (MCMC) method; details of this method are presented in the following section.

### Parameter inference using the MCMC method

In the MCMC model, there are three unknown parameters, a, b, and c, which were estimated using a robust Bayesian method [31]. The likelihood function of the influenza observations, Y, is defined as follows:

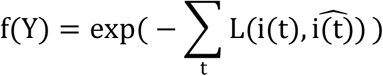

where 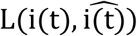 is the loss from using 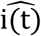 to estimate i(t), and Y is the total i(t). Because i(t) is a small value in real observations, the squared loss function is not sufficient to capture the deviation between 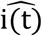 and i(t). Thus, we used the abstract loss function, i.e., 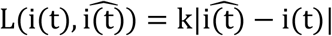, where k is a constant, to increase the accuracy; in this study, k=1000. Prior distributions for a, b, and c were determined to be normal, and M-H within the Gibbs algorithm was used to sample from posterior distributions. The estimation results for the parameters a, b, and c are reported in the supplemental information.

### Forecasts

An accurate prediction of influenza dynamics is challenging because of several factors that are unknown, such as influenza virus evaluation and climate change. All of these factors contribute to the cross-seasonal variability in influenza incidence. In this study, without considering the impact of climate change and influenza virus evaluation on influenza transmission in 2020–2025, we simply considered the impact of changes in influenza vaccination coverage before and after the COVID-19 pandemic. The influenza vaccination rate before the COVID-19 pandemic was 10.0% (95% confidence interval [CI]: 7.1%–13.4%), and that after the COVID-19 pandemic was 16.7% (95% CI: 12.9%–21.0%) [32]. We assumed that the vaccine effectiveness was 59.0% (95% CI: 51.0%–66.0%) [25]. In the 2020–2021 season, we assumed that the reproduction number of influenza was reduced by 55% [29]. Our previous study also found a similar reduction, that is, we observed that the incidence rate of influenza decreased by 64% in the following 4 weeks after NPIs were implemented on January 23, 2020 [8].

In the 2021 – 2022 epidemiological season, we further estimated the reduction in the transmission efficiency of the influenza virus based on surveillance data. Specifically, we introduced a multiplier, θ ∈ [0, 1], to represent the percentage reduction, a, caused by NPIs in the 2021 – 2022 season. We calculated the value of θ that best fit the incidence rates in the 2021–2022 epidemiological season in northern China and that in southern China.

In the 2022–2025 seasons, we predicted the influenza transmission dynamics under the following three scenarios:

Scenario 1: Complete relaxation of NPIs on December 4, 2022 and maintained until 2025, in which the transmission efficiency of the influenza virus was same to that of the value estimated from the data in the 2018–2019 and 2019–2020 seasons.

Scenario 2: Partial relaxation of NPIs in on December 4, 2022 to October 1, 2023, which led to a 10% reduction in the transmission efficiency of the influenza virus in 2022 compared with the estimated value from the data in the 2018 – 2019 and 2019 – 2020 season, and complete relaxation in 2023–2025.

Scenario 3: Partial relaxation of NPIs on December 4, 2022 to October 1, 2023, which led to a 20% reduction in the transmission efficiency of the influenza virus in 2022 compared with the estimated value from the data in the 2018 – 2019 and 2019 – 2020 seasons, and complete relaxation in 2023–2025.

Scenario 4: The transmission efficiency of the influenza virus in the year 2022 was same to that in 2021, and NPIs were completely relaxed in 2023–2025.

## Results

Since January 23, 2020, when the Chinese government began a one-level response and implemented a set of NPIs to mitigate the COVID-19 pandemic, the incidence rate of influenza decreased rapidly in both southern China and northern China (Figure 1). During the 2020–2021 season, the activity of influenza and number of new daily cases of COVID-19 remained at an extremely low level. In the 2021–2022 season, the activity of influenza rebounded in both southern China and northern China, but it still had a relatively lower peak than those in seasons before the COVID-19 pandemic. Compared with the peak incidence rates of influenza in the 2018–2019 and 2019–2020 seasons, the peak incidence rate of influenza in the 2021 – 2022 epidemiological year in southern China was 70.9% lower, whereas that in northern China was only 32.4% lower. Furthermore, in the 2021–2022 season, we estimated that the transmission efficiencies of influenza in southern China and northern China were reduced by 64.0% (95% CI: 62.8%–64.9%) and 34.5% (95% CI: 34.4%–34.7%), respectively.

**Figure 1.**
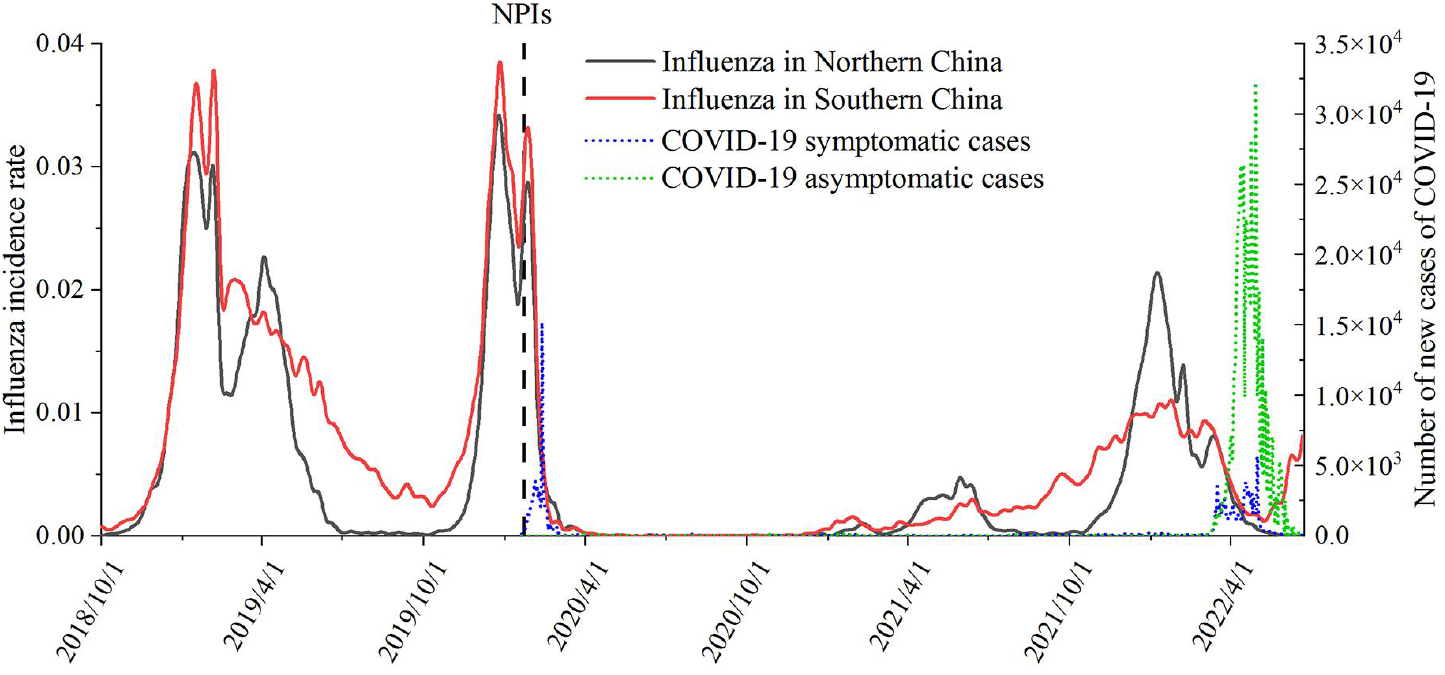
Influenza and COVID-19 activities in China from 2018–2022.

We simulated the influenza transmission from 2018 – 2020 and calibrated it using surveillance data in southern China and northern China respectively. The simulated influenza incidence rate in 2018 – 2020 matched the observed incidence rate well (R^2^=0.96 in southern China, and R^2^=0.98 in northern China) (Figure 2), regardless of whether the average duration of immunity to the influenza virus was set as 2 years or 3 years.

**Figure 2.**
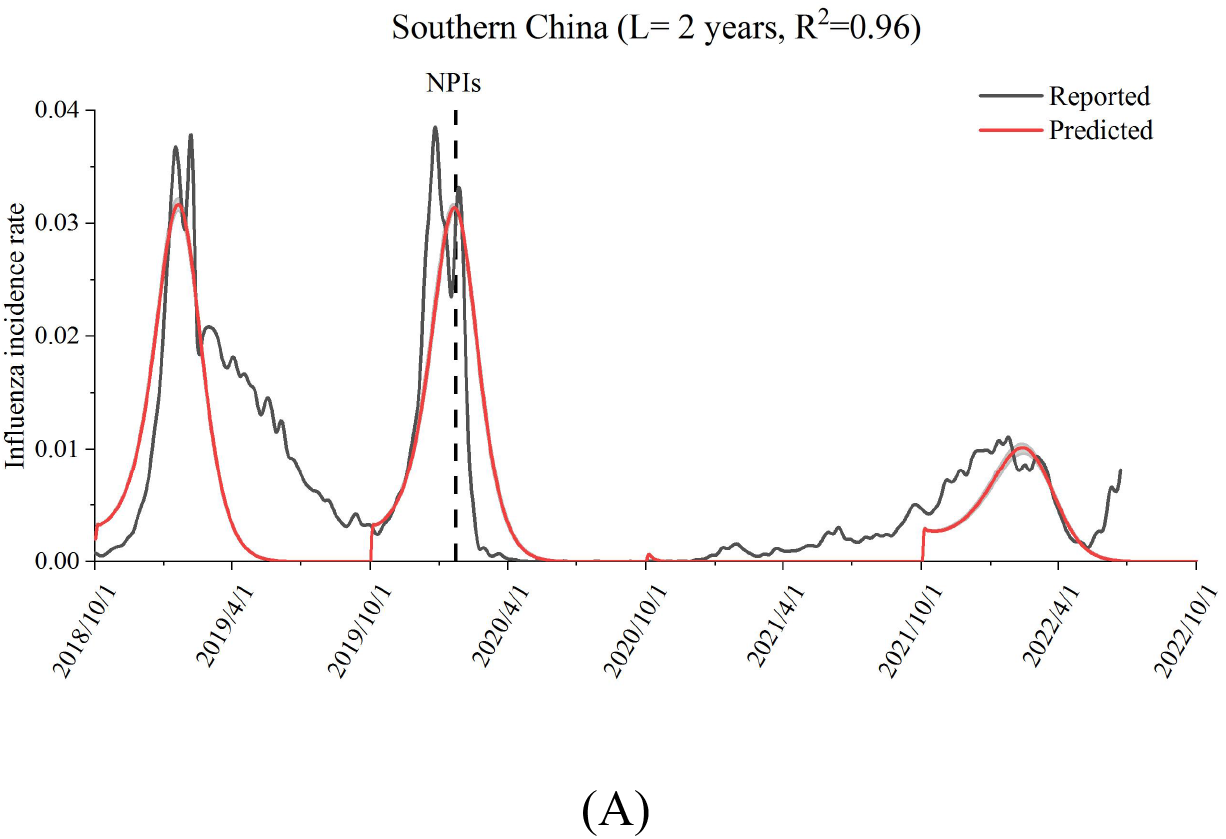
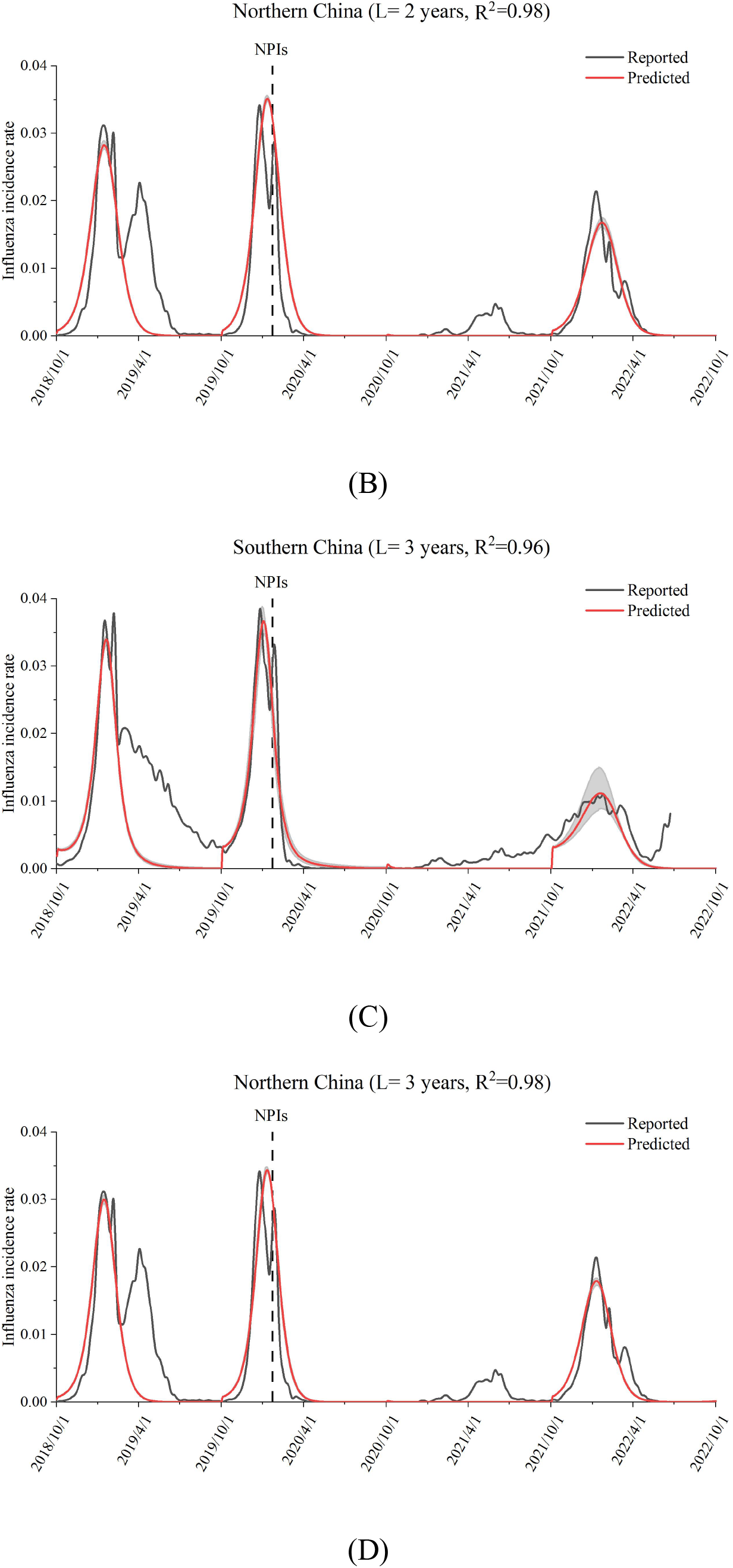
Fitted and surveillance influenza activities in 2018–2022 in southern China (A, C) and northern China (B, D). The average duration of immunity was assumed to be 2 years (A, B) or 3 years (C, D). The gray-shaded area represents the 95% confidence interval.

In the 2018– 2019 and 2019 –2020 epidemiological years, when the average immunity period was set as 2 years, the estimated percentage of susceptible individuals in the population at the beginning of the epidemiological year was approximately 45%–50%, it reached the lowest value in February, and peaked in November (Figure 3). When the immunity period was set as 3 years, the estimated percentage of individuals in the population who are susceptible to influenza was much lower, at approximately 20%– 32%. This was reasonable because, with a longer immunity period, the recovery period would also be longer (Figure 3). According to a community-based cohort study of the 2018 – 2019 epidemiological year in China, the percentage of individuals in the population who were susceptible to influenza virus was approximately 25%–37% [33]; thus, in the following study, the average immunity period was set as 3 years.

**Figure 3.**
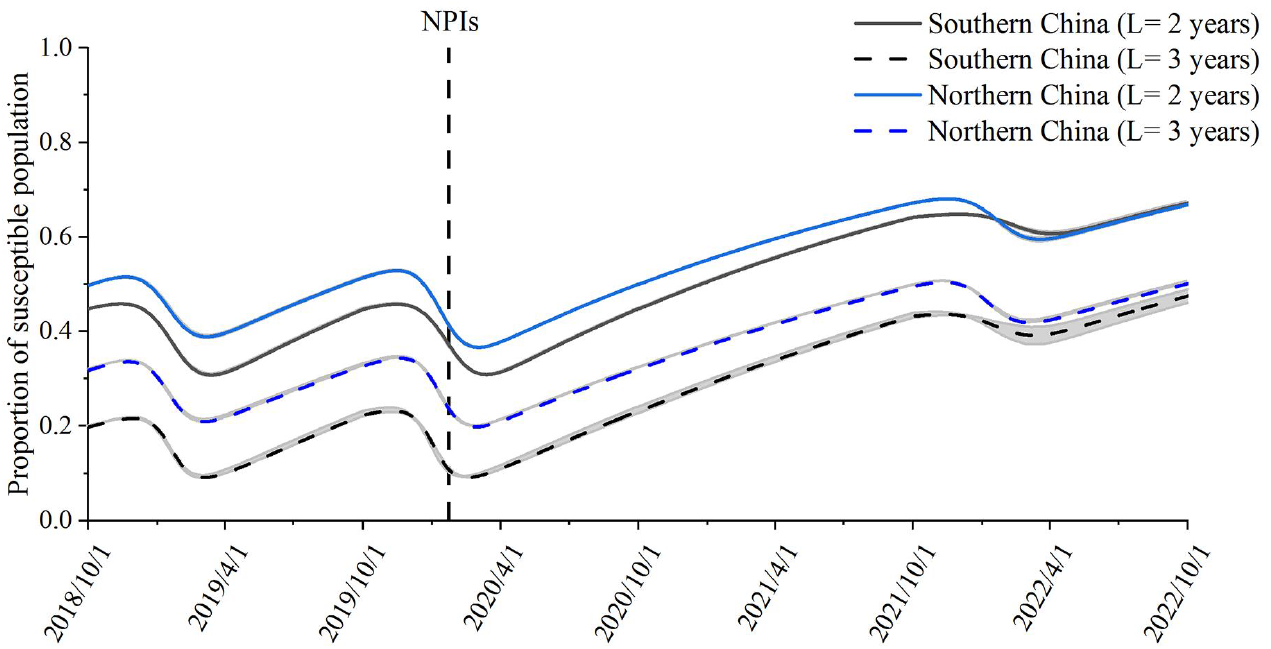
Simulated population susceptibility to influenza in southern China and northern China from 2018–2022 accumulate over time. The average duration of immunity was assumed to be 2 or 3 years. The gray-shaded area represents the 95% confidence interval.

Because the incidence rate of influenza from 2020 – 2022 was very low, it was not surprising that the accumulation of population susceptibility reached an extremely high level at the end of 2021 and remained at a high level by 2022 (Figure 3). When the immunity period was assumed to be 3 years, by the end of 2021, the percentages of influenza-susceptible individuals in the population in northern China and population in southern China increased by 57.3% and 138.6%, respectively.

The predicted transmission dynamics of influenza from 2022 – 2025 in southern China and northern China under the four different scenarios are shown in Figure 4. When the immunity period was assumed to be 3 years, the relaxation of NPIs would lead to a substantial increase in the incidence rate of influenza in the 2022–2023 epidemiological year in both southern China and northern China (Figure 4). When the NPIs were completely relaxed, the peak incidence rate of the influenza epidemic in southern China and that in northern China would increase by 3.7-fold and 3.0-fold in the 2022 – 2023 epidemiological year (Table 1). With a greater relaxation of NPIs, the rebound of influenza activity is more pronounced in the year 2022-2023 (Figure 4, Table 1). In addition, with the relaxation of NPIs on December 4, the peak influenza activity predicted in the 2022 – 2023 season would occur later than that in years before the COVID-19 pandemic, but the speed of reaching the peak value is faster. The peak timing would be around January to February in the 2022 – 2023 season, while in the 2018–2019 and 2019–2020 seasons, the peak timing was around December to February (Table 1). In Scenario 4, the rebound of influenza activity in 2023-2024 was approximately close to the rebound level of influenza activity in 2022-2023 in Scenario 1. This means that later relaxation of NPIs would not much higher rebound of influenza activities in China. The main reason may be that under recent condition, though the influenza activities is low, it is prevalent.

**Table 1.** Peak incidence rates and peak timings of the influenza epidemic in the epidemiological years 2018–2024 before and after the implementation of strict NPIs in different scenarios.

| Year | Region | Peak arrival timing |  | Peak incidence rate |
| --- | --- | --- | --- | --- |
| 2018–2019 | Northern | January 13 |  | 0.0312 |
|  | Southern | February 5 |  | 0.0379 |
| 2019–2020 | Northern | December 24 |  | 0.0324 |
|  | Southern | December 25 |  | 0.0385 |
| 2021–2022 | Northern | January 7 |  | 0.0215 |
|  | Southern | January 23 |  | 0.0111 |
| Prediction |  |  |  |  |
| 2022–2023 | Northern | Scenario 1 | February 5 | 0.1273 (+300.3%)<sup>a</sup> |
|  |  | Scenario 2 | February 14 | 0.0987 (+210.4%)<sup>a</sup> |
|  |  | Scenario 3 | February 28 | 0.0689 (+116.7%)<sup>a</sup> |
| 2023–2024 |  | Scenario 4 | December 3 | 0.1224 (+284.9%)<sup>a</sup> |
| 2022–2023 | Southern | Scenario 1 | January 12 | 0.1822 (+377.0%)<sup>a</sup> |
|  |  | Scenario 2 | January 16 | 0.1538 (+302.6%)<sup>a</sup> |
|  |  | Scenario 3 | January 23 | 0.1257 (+229.1%)<sup>a</sup> |
| 2023–2024 |  | Scenario 4 | November 9 | 0.1668 (+336.6%)<sup>a</sup> |

<sup>a</sup>Percentage increase in the peak incidence rate from the mean peak value in the 2018–2019 and 2019–2020 epidemiological years.

**Figure 4.**
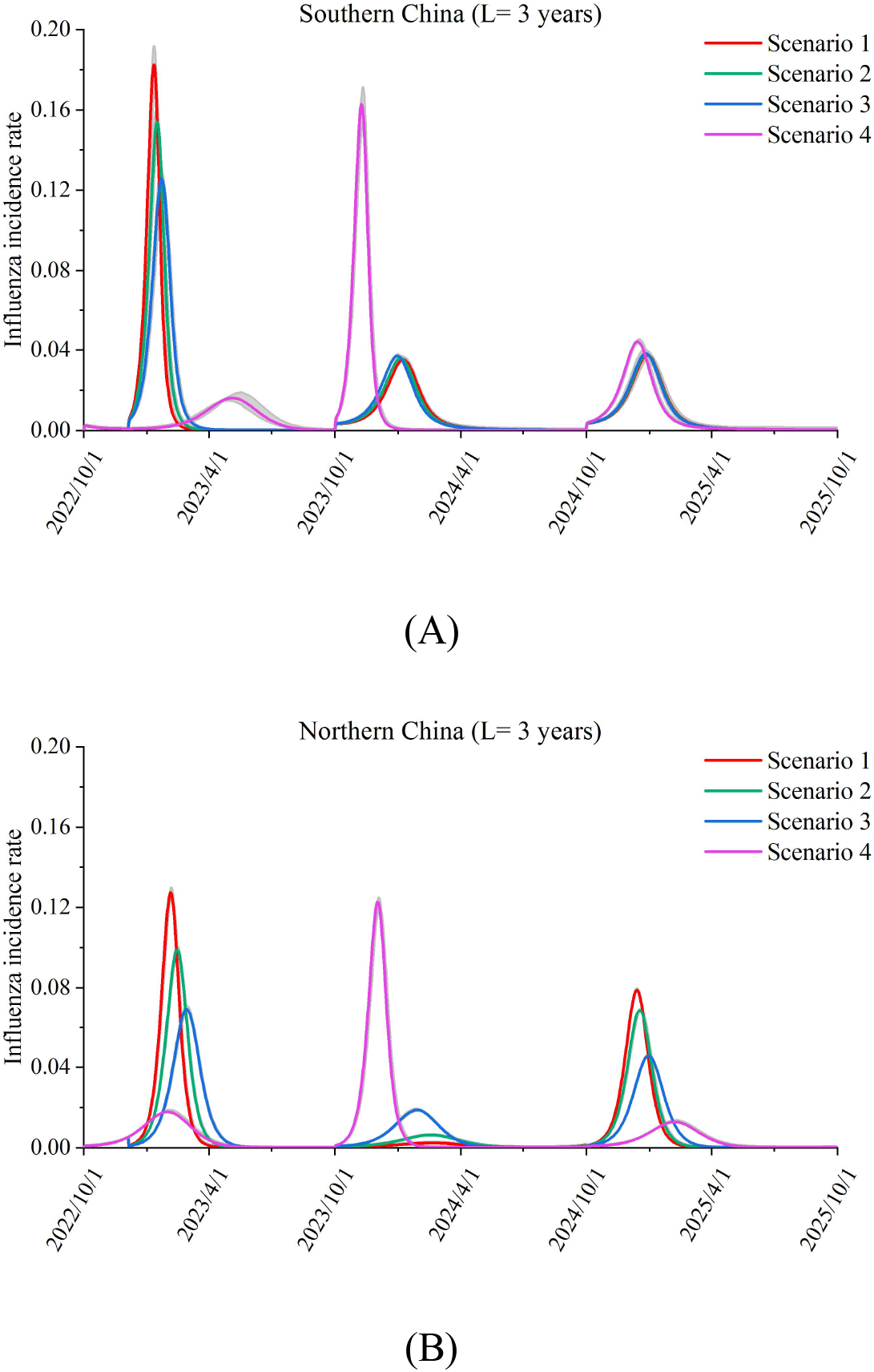
Predicted influenza activities in the 2022–2023, 2023–2024, and 2024–2025 epidemiological years in (A) southern China and (B) northern China. The average duration of immunity was assumed to be 3 years. The gray-shaded area represents the 95% confidence interval.

The impacts in southern and northern China were slightly different. The relaxation of NPIs in southern China would lead a larger rebound of the influenza epidemic than that in northern China (Figure 4). The main reason for this could be that the influenza activity in the 2021–2022 epidemiological year in southern China was lower than that in northern China (Figure 2). The relaxation of NPIs had a very limited effect on influenza activity in the 2024–2025 epidemiological years in southern China in all four scenarios; that is, the incidence rates of influenza in the 2024–2025 seasons gradually returned to the level of that in years before the COVID-19 pandemic (Figure 4A). The impact in northern China still existed in the 2023–2024 and 2024–2025 epidemiological years. In northern China, a high influenza activity in a year would lead to lower influenza activity in the subsequent year (Figure 4B).

A potential measure for controlling the rebound of the influenza epidemic when relaxing NPIs is vaccination. Figure 5 presents the prediction of the influenza incidence rate in the 2022–2023 season under different influenza vaccination coverage rates when the NPIs were completely relaxed. When the influenza vaccination coverage rates in southern China and northern China reached 53.8% (95% CI: 51.1%–55.8%) and 33.8% (95% CI: 33.5% – 34.3%) (Figure 5), respectively, the influenza incidence rates remained at the same level as those before the COVID-19 pandemic, even when the NPIs are completely relaxed.

**Figure 5.**
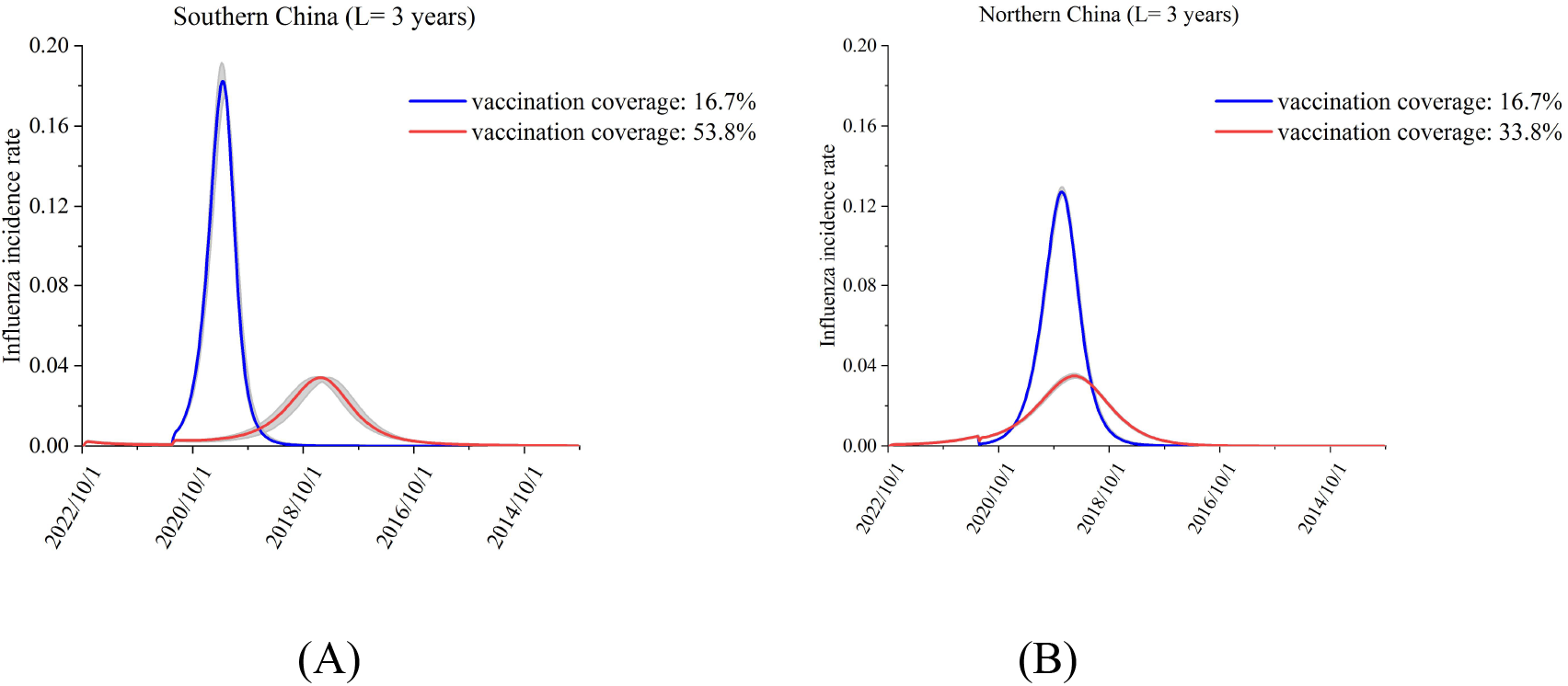
Predicted influenza activities in the 2022–2023 epidemiological year under different influenza vaccine coverage rates in (A) southern China and (B) northern China. The gray-shaded area represents the 95% confidence interval.

## Discussion

During the 2019–2020 season, the influenza activity in China was considerably reduced after NPIs for COVID-19 control were implemented [8]. In the 2020 – 2021 epidemiological year, there was almost no influenza activity. In the 2021 – 2022 epidemiological year, the influenza activity rebounded; however, the peak influenza incidence rates in southern China and northern China were still 70.9% lower and 32.4% lower, respectively, than those before the pandemic, and the transmission efficiencies of influenza in southern China and northern China were estimated to be 64.0% lower and 34.5% lower, respectively. In the short term, NPIs are expected to decrease not only the activity of influenza but also the activity of other infectious diseases [11, 34] and alleviate the burden on healthcare systems. However, after the relaxation of NPIs, the potential accumulation of susceptible individuals to influenza infection may lead to a large outbreak, which would be worrisome for the medical infrastructure if no interventions were implemented. Our modeling suggests that, in the upcoming 2022 – 2023 season, the peak influenza incidence rates in southern China and northern China will be greater than three-fold higher than those before the COVID-19 pandemic if the NPIs are completely relaxed. Moreover, the rebound in southern China would be more pronounced than that in northern China, which is partly due to the relatively low rebound of influenza activity in the 2021–2022 epidemiological year in southern China. The higher increase in the proportion of susceptible people in southern China than that in northern China led to a higher influenza rebound in southern China, which is consistent with the results of other studies [35]. In addition, if NPIs were lifted in the 2022 – 2023 epidemiological year, the influenza activity may have arrived faster. The rebound of influenza activity after lifting NPIs has been reported in several countries in which NPIs were partially or completely relaxed during the 2021 – 2022 season [18]. However, in China, the NPIs were not completely lifted in the 2021 – 2022 epidemiological year, so there was a relatively small rebound of influenza activity in the winter and spring months. Thus, compared with other countries, China may exhibit a greater accumulation of individuals in the population who are susceptible to influenza, leading to a larger rebound in the influenza activity after relaxing NPIs.

Vaccination for influenza could be an important intervention to control the rebound of influenza activity after the COVID-19 pandemic in China. As the rate of influenza vaccination was low before the COVID-19 pandemic [36], improving the coverage of influenza vaccination in China has become particularly important. Our study suggests that, when the influenza vaccination rate increases to 54% and the vaccine effectiveness is 59%, even with the complete relaxation of NPIs, the influenza activity could remain at a pre-pandemic level in the 2022–2023 season. In addition, we also demonstrated that southern China needs a higher vaccination rate (54%) than northern China (34%) to achieve the same outcome, which is consistent with the results of the study by Ali et al. [35]. However, owing to the low prevalence of influenza activity during the past 2 years, it may be difficult to predict the dominant influenza strains to guide influenza vaccine production. Before the COVID-19 pandemic, in the 2018 – 2019 and 2019 – 2020 epidemiological years, the incidence of influenza was dominated by that of the influenza A sub-type, whereas after the COVID-19 pandemic, it was dominated by the influenza B Victoria sub-type [37]. Thus, the uncertainty of the dominant influenza strains after the pandemic will decrease the effectiveness of influenza vaccination. Therefore, improving the coverage of influenza vaccination by promoting a reimbursement-based influenza vaccination policy [38] and implementing some NPIs, such as wearing masks, at the same time may be a better option.

Several studies have quantified the rebound of influenza activity in the coming season, especially with NPIs being completely relaxed, including studies in China, the United Kingdom, and the United States [29, 39]. It is not surprising that these studies demonstrated that the severity of influenza rebound was related to the duration of the implementation of NPIs and various relaxations of NPIs. The longer the duration of NPIs implementation is and the more thoroughly relaxed NPIs are, the more serious the influenza rebound is.

Some limitations of this study must be noted, as they may affect the generalization of the results. First, in addition to the NPIs and influenza vaccination, the transmission dynamics of the influenza epidemic can be affected by viral antigenic evolution [40], climatic conditions [41], and host (e.g., human) contact patterns [42]. We did not consider these factors in the present study. Second, influenza and SARS-CoV-2 co-infection has been reported in experimental data [43], and the interaction between these viruses interferes with the level of immunity of the population [44]. Our study did not consider the cross-protection of SARS-CoV-2 against infection by the influenza virus. In the future, we will strive to obtain population-level immunity data to predict influenza activity with a higher accuracy. Third, the reported influenza incidence rate in the past 2 years may be biased. Owing to the impact of the COVID-19 pandemic, some patients with influenza may prefer to stay at home and avoid going to the hospital. Therefore, the incidence rate of influenza during this period may be an underestimation of the true incidence rate, leading to a biased estimation of the proportion of susceptible individuals. Finally, we failed to retrieve age-specific influenza incidence data, so we could not predict the influenza incidence trend in different age groups in the next 3 years or provide insightful opinions. We will strive to obtain age-specific influenza incidence data and provide suggestions for age-stratified target-based vaccination strategies and other effective prevention and control measures.

## Conclusions

In conclusion, this study quantified the rebound of influenza activity in southern China and northern China in the 2023–2024 and 2024–2025 epidemiological years when NPIs are relaxed. We demonstrated that the greater the extent of the relaxation of NPIs was and the longer the period of immunity to the influenza virus was, the larger the rebound of influenza activity was and the faster the arrival of influenza activity was. The peak incidence rate of influenza could greater than three-fold than that before the pandemic, and the rebound of influenza activity in southern China would be larger than that in northern China. This finding is mainly due to the accumulation of individuals in the population who are susceptible to influenza. Therefore, influenza vaccination should be advocated to reduce the reemergence of influenza. To control the influenza epidemic to the pre-pandemic level after completely relaxing NPIs, the influenza vaccination rates in southern China and northern China should increase to 53.8% and 33.8%, respectively.

## Supporting information

Supplemental information includes two figures.

## Data Availability

All data produced in the present study are available upon reasonable request to the authors

## Description of supplemental information

Supplemental information includes two figures.

## Acknowledgments

YS, DW, SY and HL conceived, designed and supervised the study. HL and MY analyzed the data. MY performed the simulation. YL, JT, JY, MT and DW collected the data. HL and YL cleaned data. HL and MY wrote the drafts of the manuscript. YS, DW, SY and LY commented on and revised drafts of the manuscript. All authors read and approved the final report. This study was funded by grants from the National Natural Science Foundation of China (Grant No. 82003509, 82173577, 81672005), and the Natural Science Foundation of Zhejiang Province (Grant No. LQ20H260009).

