## Supplemental information includes two figures. for "Quantifying the rebound of influenza epidemics after relaxing nonpharmaceutical interventions during the coronavirus disease 2019 pandemic in China"

### Parameter inference

When w = 0.017, D = 5 days and L = 2 years, the best fitting parameters [ (mean and 95% confidence intervals (*CI*)] in Southern China in 2018-2020 season were determined to be a = 0.43072 (95% *CI*: 0.42867, 0.43292); b = 0.29728 (95% CI: 0.29162, 0.30000); c = 0.13932 (95% *CI*: 0.11905, 0.16003) and the best fitting parameters in Northern China in 2018-2020 season were a = 0.41373 (95% CI: 0.41176, 0.41555); b = 0.29841 (95% CI: 0.29531, 0.30000); c = 0.94879 (95% CI: 0.92021, 0.97527) (Figure S1A, B).

When w = 0.017, D = 5 days and L = 3 years, the best fitting parameters in Southern China in 2018-2020 season were determined to be a = 1.14922 (95% CI: 1.07660, 1.20520); b = 0.26190 (95% CI: 0.24162, 0.29060); c = -0.92742 (95% CI: -1.12812, -0.68641) and the best fitting parameters in Northern China in 2018-2020 season were a = 0.63986 (95% CI: 0.63679, 0.64345); b = 0.29726 (95% CI: (0.29092, 0.30000); c = 0.58773 (95% CI: 0.56141, 0.61308) (Figure S1C, D).

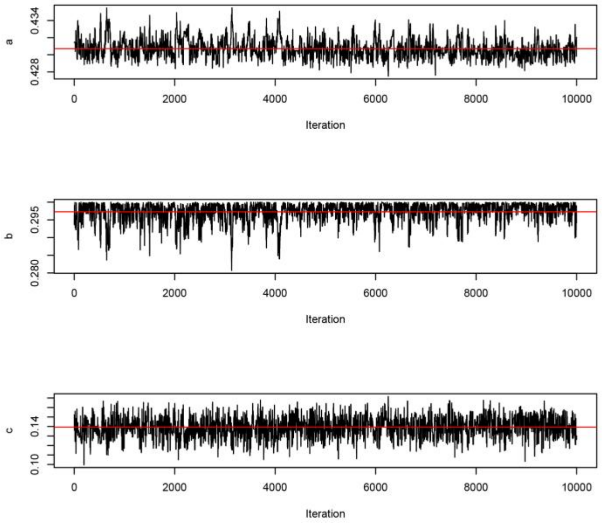


(A)

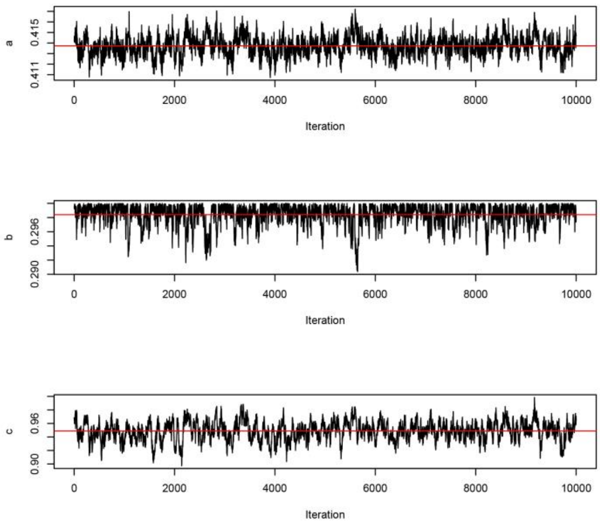
a
0.411
0.415
b
0.290
0.296
c
0.90
0.96
0
2000
4000
6000
8000
10000
Iteration

(B)

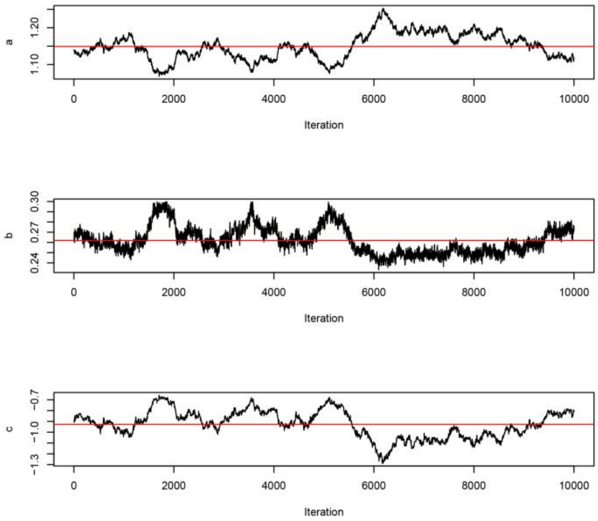
a
1.10
1.20
b
0.24
0.27
0.30
c
−1.3
−1.0
−0.7
0
2000
4000
6000
8000
10000
Iteration

(C)

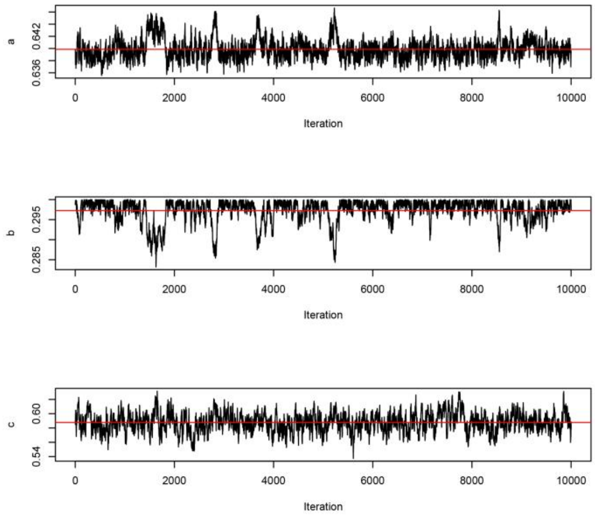
a
0.636
0.642
b
0.285
0.295
c
0.54
0.60
0
2000
4000
6000
8000
10000
Iteration

(D)

**Figure S1**. Monte Carlo Markov Chain sampling of a, b and c, with w=0.017, D=5 days and L=2 (A, B) and 3 (C, D) years in Southern (A, C) and Northern China (B, D) in 2018-2020.

When w = 0.017, D = 5 days, L = 2 years, b = 0.29728 and c = 0.13932, the best fitting parameters of a in Southern China in 2021-2022 season was 0.27064 (95% CI: 0.26998, 0.27124). When w = 0.017, D = 5 days, L = 2 years, b = 0.29841 and c = 0.94879, the best fitting parameters of a in Northern China in 2021-2022 season was 0.29752 (95% CI: 0.29608, 0.29904) (Figure S2A, B).

When w = 0.017, D = 5 days, L = 3 years and b = 0.26190, the best fitting parameters of a, c in Southern China in 2021-2022 season were a = 0.41339 (95% CI:0.41221 0.41460), c = 0.48888 (95% CI: 0.37802, 0.57838). And when w = 0.017, D = 5 days, L = 3 years and b = 0.29726, the best fitting parameters of a, c in Northern China in 2021-2022 season were a = 0.41928 (95% CI: 0.41723, 0.42124), c = 1.22282 (95% CI:1.18068, 1.26087) (Figure S2C, D).

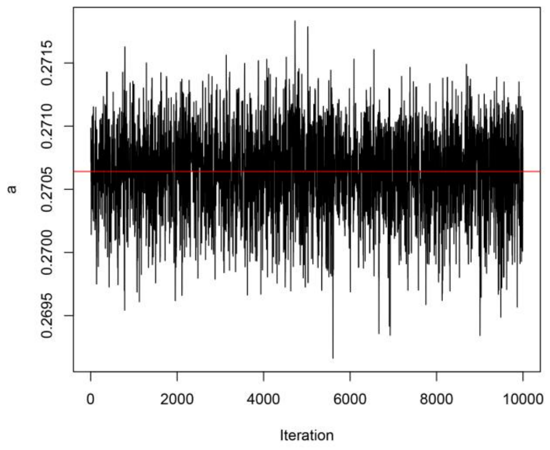

0.2695
0.2700
0.2705
0.2710
0.2715
a
0
2000
4000
6000
8000
10000
Iteration


(A)

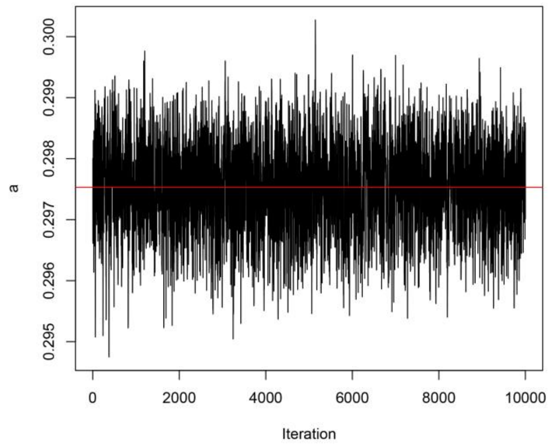

0.295
0.296
0.297
0.298
0.299
0.300
a
0
2000
4000
6000
8000
10000
Iteration


(B)

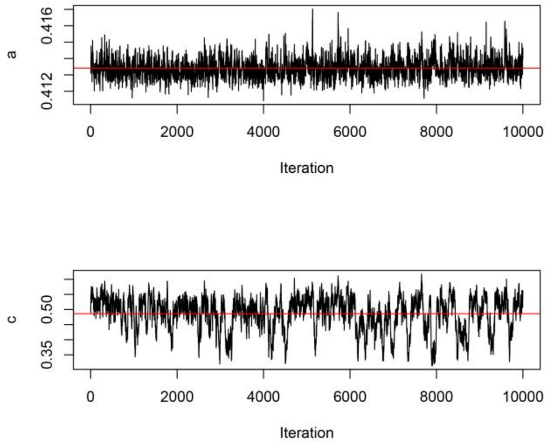

0.412
0.416
a
0
2000
4000
6000
8000
10000
Iteration
0.35
0.50
c
0
2000
4000
6000
8000
10000
Iteration


(C)

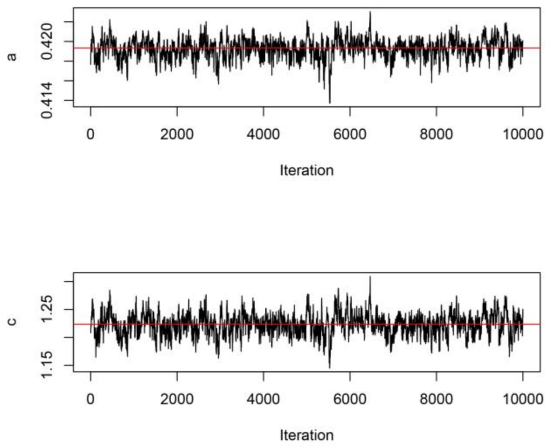


(D)

**Figure S2.** Monte Carlo Markov Chain sampling of a, with L=2 (A, B) and 3 (C, D) years in Southern (A, C) and Northern China (B, D) in 2021-2022.
